# Perceived and Enacted HIV-Related Stigma in Eastern and Southern Sierra Leone: A Psychometric Study of an HIV Stigma Scale

**DOI:** 10.1101/2023.07.07.23292152

**Authors:** George A. Yendewa, Rashid A. Kpaka, Edmond Sellu, Sahr A. Yendewa, Samuel P.E. Massaquoi, Peterlyn E. Cummings, Manal Ghazzawi, Umaru Barrie, Karine Dubé, Sulaiman Lakoh, Peter B. James, Robert A. Salata, Lawrence S. Babawo

**Affiliations:** Department of Medicine, Case Western Reserve University School of Medicine, Cleveland, Ohio, USA; Division of Infectious Diseases and HIV Medicine, University Hospitals Cleveland Medical Center, Cleveland, Ohio, USA; Johns Hopkins Bloomberg School of Public Health, Baltimore, Maryland, USA; Eastern Technical University, Kenema, Sierra Leone; Department of Nursing, School of Community Health Sciences, Njala University, Bo Campus, Sierra Leone; Ministry of Health and Sanitation, Freetown, Sierra Leone; KnowHep Foundation, Freetown, Sierra Leone; University of Texas Southwestern Medical Center, Dallas, Texas, USA; Division of Infectious Diseases and Global Public Health, School of Medicine, University of California San Diego, La Jolla, California, USA; AntiViral Research Center, University of California San Diego, San Diego, California, USA; Health Policy and Management, UNC Gillings School of Global Public Health, Chapel Hill, North Carolina, USA; Department of Medicine, College of Medicine and Allied Health Sciences, University of Sierra Leone, Freetown, Sierra Leone; Connaught Hospital, University of Sierra Leone Teaching Hospitals Complex, Freetown, Sierra Leone; Faculty of Health, Southern Cross University, Lismore, Australia

**Keywords:** HIV, stigma, discrimination, mental health, Sierra Leone

## Abstract

**Background:** HIV stigma continues to hinder the care of people with HIV (PWH), especially in low-resource settings. We aimed to adapt and validate a concise HIV stigma scale for perceived HIV stigma in Sierra Leone.

**Methods:** We enrolled participants in two HIV clinics in Eastern and Southern Sierra Leone in 2022. We assessed perceived stigma using a 12-item adaptation of Berger’s HIV Stigma Scale and enacted stigma using select USAID indicators. We used ordinal logistic regression to identify predictors of perceived stigma and Pearson’s correlation to examine associations between perceived and enacted stigma.

**Results:** 624 PWH were enrolled. The final adapted 6-item HIV stigma scale demonstrated acceptable internal consistency (Cronbach’s α = 0.72) and a four-factor solution accounting for 84.8% of variance: concern about public attitude (2 items), personalized stigma (2 items), negative self-image (1 item), and disclosure concerns (1 item). The prevalence of perceived HIV stigma was 68.6%, with disclosure concerns as the most prominent contributor. Enacted HIV stigma was reported by only 6.7% of participants, with partner/spousal abandonment and workplace stigma being the most common discriminatory experiences. Employment (β = 0.525, p <0.001), residence in Eastern Sierra Leone (β = 3.215, p < 0.001), and experiencing enacted stigma (β = 0.804, p < 0.001) were significantly associated with perceived stigma. Having a family member or friend with HIV (β = -0.499, p < 0.001), and HIV disclosure (β = -0.710, p < 0.001) were protective against perceived stigma. Enacted stigma strongly correlated with partner abandonment and family isolation (r = 0.223, p < 0.001).

**Conclusion:** We found high levels of perceived HIV stigma, underscoring the need for targeted interventions to combat stigma and promote inclusivity for PWH in Sierra Leone.

## INTRODUCTION

The epidemic associated with the human immunodeficiency virus (HIV) continues to present a pressing public health problem. Globally, approximately 38 million people were living with HIV (PWH) in 2021, with the majority (67%) residing in the World Health Organization (WHO) African region [1]. Although there is currently no cure for HIV, the widespread availability of combination antiretroviral therapy (ART) has significantly improved the survival of PWH [1, 2]. However, despite the success of ART in prevention and treatment, HIV-related stigma and discrimination persist, posing a formidable barrier in the global response to the HIV epidemic.

Stigma has been defined as *a set of attributes that is deeply discrediting* [3]. As a complex social phenomenon, stigma encompasses negative attitudes, beliefs, and behaviors directed towards individuals or groups based on perceived differences or characteristics that deviate from societal norms [3-5]. HIV-related stigma is multifaceted in its expression. *Perceived* (also referred to as *felt*) stigma refers to the subjective beliefs and attitudes held by PWH about the negative judgments and social rejection they anticipate from others [6]. *Enacted* stigma, on the other hand, encompasses the actual experiences of discrimination, mistreatment, and marginalization faced by PWH due to their real or perceived serostatus [6]. HIV stigma can have profound impacts on the psychosocial well-being, mental health, quality of life, and clinical outcomes of PWH [7-17]. Studies have shown that HIV stigma leads to increased levels of depression and anxiety [7], unwillingness to test for HIV [8, 9], fear of status disclosure [10, 11], and poor treatment adherence [12, 13]. Furthermore, the fear of stigma contributes to poor retention in HIV care, compromising treatment outcomes [14]. Moreover, HIV stigma is linked to higher HIV viremia and lower CD4 cell counts, indicating negative effects on disease progression and immune health [15-17]. Thus, understanding and addressing the various dimensions of HIV stigma is essential for comprehensive approaches to HIV prevention, care, and psychosocial support.

In Sierra Leone, the HIV epidemic remains a concern, with 1.7% of the population estimated to be living with HIV [18]. Only a handful of studies have documented HIV stigma in Sierra Leone. A study by Kelly et al. [9] found that HIV stigma had an inverse association with HIV testing at both the individual and community levels in Sierra Leone. The negative association between HIV stigma and testing was stronger at the community level compared to the individual level [9]. Another study by Kelly et al. [20] demonstrated that HIV stigma at the community level significantly influenced disclosure concerns, condom usage, and self-reported sexually transmitted diseases, especially among women. Additionally, a qualitative study by Lahai et al. [20], involving 16 PWH and 4 healthcare workers identified stigma as a major barrier to ART adherence. These findings underscore the exigent need for targeted interventions addressing HIV stigma at both individual and community levels in Sierra Leone. However, to the best of our knowledge, no study to date has measured HIV stigma from the perspective of PWH in the country. This could be due, in part, to the lack of validated instruments to measure HIV-related stigma in this setting.

Our study had two main objectives: (1) to adapt and validate a concise HIV stigma scale, and (2) to assess the levels of perceived and enacted HIV stigma from the perspective of PWH in Sierra Leone.

## METHODS

### Participants, study sites and recruitment

We employed convenience sampling to consecutively enroll participants in two hospital-based HIV treatment clinics in Sierra Leone between August and November 2022. Bo Government Hospital (BGH) is a 400-bed secondary health facility that serves over 1 million individuals in the Southern region. Kenema Government Hospital (KGH) is a regional 350-bed hospital providing health services to 670,000 people in the Eastern region of Sierra Leone.

Trained research staff approached potential study participants at the HIV treatment clinics in BHG and KGH and provided them with information about the study. Those who expressed interest and provided informed consent were enrolled in the study. The inclusion criteria were age ≥ 18 years, confirmed positive HIV serostatus by an approved diagnostic method such as serological testing or polymerase chain reaction methods, or documentation of infection in clinic records. Exclusion criteria included age < 18 years and being unable or unwilling to provide informed consent.

The research staff were native Sierra Leoneans with experience in survey methods and a deep understanding of the local context, including languages, norms, and customs. They were affiliated with Njala University in the Southern region and Eastern Technical University in Kenema, both public tertiary educational institutions in Sierra Leone. Before the study, a one-week training seminar was conducted to discuss and make necessary modifications to the survey methods and instruments based on feedback from researchers.

### HIV stigma assessment tools and procedures

We collected socio-demographic information (sex, age, relationship status, educational attainment, employment, and religion) and HIV-related history (HIV status disclosure, duration since diagnosis, and having family or friends living with HIV) using a survey questionnaire.

We assessed enacted HIV stigma using four questions adapted from the United States Agency for International Development (USAID) indicators for assessing discrimination toward PWH [21]. The indicators assessed partner/spousal abandonment, isolation from family members, social exclusion, and workplace stigma [21]. Participants responded with “yes” or “no” to indicate whether they had experienced each form of enacted stigma. Each “yes” response was scored as 1 point, while a “no” response received no points. The possible range of scores for enacted HIV stigma was 0-4.

We evaluated perceived HIV stigma using the 12-item HIV Stigma Scale by Reinius et al. [22] (Supplementary Materials). This is an abridged and validated version of the 40-item HIV Stigma Scale originally developed and validated by Berger et al [23]. Similar to the full-length Berger scale, the abridged version by Reinius et al. [22] consists of four domains or subscales, with three items in each: (1) *personalized stigma* (consequences of others knowing about one’s HIV status), (2) *disclosure concerns* (worries about disclosing HIV status), (3) *concern with public attitudes* (perception of discriminatory attitudes from the public), and (4) *negative self-image* (also known as *self-stigma* or *internalized stigma*). All items on the scale (consecutively labeled Q1 to Q12) were positively worded and rated on a 4-point Likert scale with equidistant scores, as follows: 1 = strongly disagree, 2 = disagree, 3 = agree, 4 = strongly agree, with higher scores indicating higher levels of perceived HIV stigma. The possible range of scores was 12-48.

For the purposes of our study, we examined and adapted the 12-item HIV Stigma Scale by Reinius et al. [22] for *content* and *construct validity*. The scale replicated Berger’s original scale’s 4-dimensional structure and demonstrated high internal consistency (Cronbach’s α = 0.80-0.88) [18]. To assist participants without a satisfactory command of the English language, we translated the scale into Krio, the *lingua franca* and *de facto* national language, which we piloted for clarity and cultural appropriateness (Supplementary Materials). We assessed *criterion-related validity* by examining the correlation between enacted stigma and perceived stigma, as well as the influence of having family/friends with HIV on overall perceived stigma [24]. We previously validated the abridged scale for measuring hepatitis B-related stigma in Sierra Leone [25]. Moreover, the full-length Berger HIV scale has previously been validated in sub-Saharan Africa [26, 27].

### Statistical analysis

We performed statistical analysis in SPSS Version 29.0 (Armonk, NY, USA; IBM Corp). We used descriptive statistics to summarize participant characteristics. Categorical variables were reported as counts (percentages) and continuous variables as medians (interquartile ranges, IQR). We assessed the psychometric properties of the adapted 12-item HBV Stigma Scale by estimating its internal consistency (reliability), with a coefficient of Cronbach’s α > 0.70 regarded as acceptable. We performed exploratory factor analysis using principal axis factoring with orthogonal (Varimax) rotation to assess the dimensional structure of the scale. We performed exploratory factor analysis using principal axis factoring with Varimax rotation to evaluate the dimensional structure of the scale and considered eigenvalues > 1 and loading saturations > 0.3 for factor retention. Adequacy of sampling was estimated using the criteria Kaiser-Meyer-Olkin (KMO) index > 0.6 and statistically significant Bartlett’s Test of Sphericity (BTS). We used proportional-odds (ordinal) logistic regression models to identify factors associated with perceived HIV stigma. Only covariates with p <0.05 in the univariate analysis were included in the multivariate model. The correlations between domains were estimated by Pearson’s correlation coefficient (r). Statistical significance for associations was set at p < 0.05 in all analyses.

### Ethical approval

We obtained ethical approval from the Njala University Institutional Review Board (IRB) (approval date 11 June 2022). We obtained written informed consent from all participants before enrolment into the study.

## RESULTS

### Study participants

We received responses from 624 PWH (100% response rate), of which 55.6% (347/624) were from Kenema (Eastern Province) and 44.4% (277/624) were from Bo (Southern Province). Most participants (71.5%, 446/624) were female and the median age was 34 years (IQR 28-42). Nearly half were married (46.2%, 288/624) and the majority were unemployed (56.4%, 352/624) and Muslim (51.9%, 324/624). About 35.3% (220/624) received no formal education. The median duration from HIV diagnosis was 5 years (IQR 3-9) and most were on treatment (96.2%, 600/624). The self-reported HIV disclosure rate was high (59.6%, 372/624), while 23.7% (148/624) reported having a family member or friend living with HIV. Table 1 summarizes the baseline socio-demographic characteristics of study participants.

**Table 1.** Socio-demographic and health characteristics of participants (N=624) (Sierra Leone, 2022)

| Characteristics | N (%) |
| --- | --- |
| **Gender** |  |
| Male | 178 (28.5) |
| Female | 446 (71.5) |
| **Age, years** |  |
| Median (IQR) | 34 (28-42) |
| <25 | 90 (14.4) |
| 25-34 | 233 (37.3) |
| 35-44 | 184 (29.5) |
| 45-54 | 83 (13.3) |
| ≥55 | 34 (5.4) |
| **Relationship status** |  |
| Single | 226 (36.2) |
| Married | 288 (46.2) |
| Divorced/widowed | 110 (17.6) |
| **Highest education attained** |  |
| None | 220 (35.3) |
| Primary | 149 (23.9) |
| Secondary | 172 (27.6) |
| Tertiary | 83 (13.3) |
| **Occupation** |  |
| Unemployed | 352 (56.4) |
| Informal | 226 (36.2) |
| Formal | 46 (7.4) |
| **Religion** |  |
| Christian | 300 (48.1) |
| Muslim | 324 (51.9) |
| **Residence** |  |
| Kenema (Eastern Province) | 347 (55.6) |
| Bo (Southern Province) | 277 (44.4) |
| **Time living with HIV** |  |
| Median (IQR) | 5 (3-9) |
| <5 | 273 (43.8) |
| 5-9 | 228 (36.5) |
| 10+ | 123 (19.7) |
| **On antiretroviral therapy** |  |
| Yes | 600 (96.2) |
| No | 24 (3.8) |
| **Disclosed HIV status** |  |
| Yes | 372 (59.6) |
| No | 252 (40.4) |
| **Has family member or friend with HIV** |  |
| Yes | 148 (23.7) |
| No | 252 (76.3) |

### Psychometric properties of the HIV Stigma Scale

We initially assessed the internal consistency of all 12 items (Q1 to Q12), which yielded an overall Cronbach’s α of 0.59. After removing 6 items (i.e., Q3, Q4, Q6, Q8, Q10, and Q12), the internal consistency of the remaining 6 items improved, with overall Cronbach’s α = 0.72. Exploratory factor analysis using Varimax rotation and principal axis factoring maintained the original 4-dimensional factor solution, which accounted for 84.8% of variance. Adequacy of sampling was confirmed, with Kaiser-Meyer-Olkin (KMO) index = 0.754 and conditions for Bartlett’s Test of Sphericity (BTS) satisfied (χ^2^ = 726.0, df = 15, p < 0.001). The dimensional organization of the shortened 6-item HIV Stigma Scale was as follows: (1) concern about public attitudes—2 items, (2) personalized stigma—2 items, (3) negative self-image—1 item, and (4) disclosure concerns—1 item (Table 2).

**Table 2.** Factor loadings from exploratory factor analysis of HIV Stigma Scale

| Item | Statement | Factor |  |  |  |
| --- | --- | --- | --- | --- | --- |
|  |  | 1 | 2 | 3 | 4 |
| **Q9** | Most people are uncomfortable around someone with HIV | **0.58** |  |  |  |
| **Q7** | People with HIV are treated like outcasts or rejects | **0.55** |  | 0.42 |  |
| **Q1** | Some people avoid touching me once they know I have HIV | 0.48 | **0.61** |  |  |
| **Q2** | People I care about stopped talking to me after learning I have HIV |  | **0.59** |  |  |
| **Q11** | People's attitudes about HIV make me feel worse about myself |  |  | **0.49** |  |
| **Q5** | I work hard to keep my HIV a secret |  |  |  | **0.42** |
| **Eigenvalue** |  | 2.6 | 1.5 | 1.2 | 1.0 |
| **Cumulative Percent of Variance** |  | 41.8% | 59.6% | 73.4% | 84.8% |
| **Kaiser-Meyer-Olkin Index of Sampling Adequacy** |  | KMO = 0.754 |  |  |  |
| **Bartlett's Test of Sphericity** | | $\chi^2 = 726$, df = 15, p < 0.001 | | | |

### Prevalence of perceived and enacted HIV stigma

Table 3 shows the distribution of perceived and enacted HIV stigma among PWH. The range of possible scores for the shortened 6-item HIV Stigma Scale was 6-24. The mean perceived HIV stigma score was 16.46 ± 2.92. Overall, perceived HIV stigma was prevalent in 68.6% (n=428) of the study participants. Across subscales, disclosure concerns were the most important contributor to perceived HIV stigma (84.3%, mean score 3.37 ± 0.63). Concerns with public attitudes (71.5%, mean score 5.72 ± 1.33) and negative self-image (71.0%, mean score 2.84 ± 0.89) were also major contributors, while personalized stigma contributed the least (56.2%, mean score 4.53 ± 1.21). However, only 6.7% (n=42) reported at least one instance of enacted stigma (overall mean score 0.266 ± 0.653). Abandonment by spouse/partner (9.1%) and loss of resources (7.2%) were the most common discriminatory experiences reported, followed by exclusion from social events (5.3%) and feeling isolated from family members (5.0%).

**Table 3.** Overall and domain levels of perceived and enacted HIV stigma Yendewa et al 2023

| Variables | Expected Range of HIV Stigma Score | Stigma Score | % With Stigma<sup>a</sup> |
| --- | --- | --- | --- |
| **Perceived (Felt) Stigma** |  |  |  |
| Overall (Mean ± SD) | 6-24 | 16.46 ± 2.92 | 68.6 |
| Personalized stigma | 2-8 | 4.53 ± 1.21 | 56.2 |
| Disclosure concerns | 1-4 | 3.37 ± 0.63 | 84.3 |
| Concerns with public attitudes | 2-8 | 5.72 ± 1.33 | 71.5 |
| Negative self-image | 1-4 | 2.84 ± 0.89 | 71.0 |
| **Enacted Stigma (Discrimination)** |  |  |  |
| Overall (Mean ± SD) | 0-4 | 0.266 ± 0.653 | 6.7 |
| Spousal abandonment | 0-1 | 0.091 ± 0.288 | 9.1 |
| Family isolation | 0-1 | 0.050 ± 0.217 | 5.0 |
| Social exclusion | 0-1 | 0.053 ± 0.224 | 5.3 |
| Workplace stigma | 0-1 | 0.072 ± 0.259 | 7.2 |

SD, standard deviation; <sup>a</sup> Percentage with stigma = mean score ÷ upper limit of expected score range × 100. The higher the score, the higher the level of stigma.

### Factors associated with perceived HIV stigma

Table 4 describes the proportional-odds (ordinal) univariate and multivariate logistic regression model of perceived and enacted HIV stigma and associated factors. Higher levels of perceived HIV stigma were significantly associated with being employed (β = 0.525, p < 0.001), residence in Kenema (Eastern Province) (β = 3.215, p < 0.001), and having experienced enacted stigma (β = 0.804, p < 0.001). Factors that were protective against perceived HIV stigma included a longer duration of time since HIV diagnosis (β = -0.008, p = 0.006), having a family member or friend with HIV (β = -0.499, p < 0.001), and having disclosed HIV status (β = -0.0.710, p < 0.001).

**Table 4.**
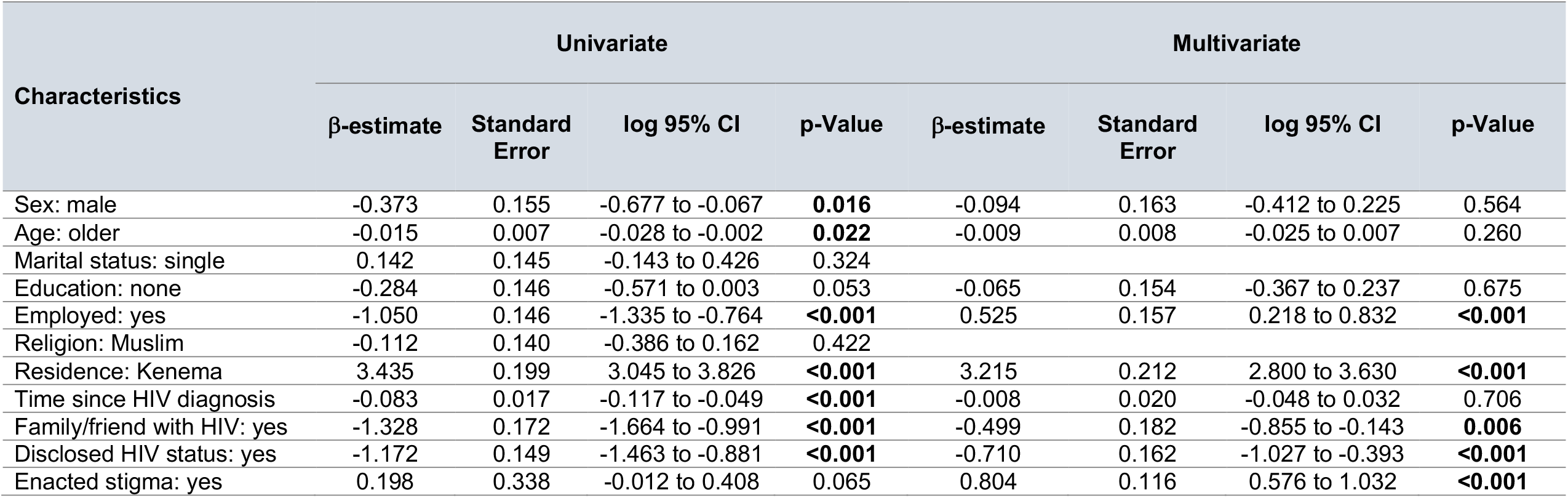
Proportional-odds (ordinal) logistic regression model of factors associated with perceived HIV stigma Yendewa et al 2023

| Characteristics | Univariate |  |  |  | Multivariate |  |  |  |
| --- | --- | --- | --- | --- | --- | --- | --- | --- |
| | β-estimate | Standard Error | log 95% CI | p-Value | β-estimate | Standard Error | log 95% CI | p-Value |
| Sex: male | -0.373 | 0.155 | -0.677 to -0.067 | **0.016** | -0.094 | 0.163 | -0.412 to 0.225 | 0.564 |
| Age: older | -0.015 | 0.007 | -0.028 to -0.002 | **0.022** | -0.009 | 0.008 | -0.025 to 0.007 | 0.260 |
| Marital status: single | 0.142 | 0.145 | -0.143 to 0.426 | 0.324 |  |  |  |  |
| Education: none | -0.284 | 0.146 | -0.571 to 0.003 | 0.053 | -0.065 | 0.154 | -0.367 to 0.237 | 0.675 |
| Employed: yes | -1.050 | 0.146 | -1.335 to -0.764 | **<0.001** | 0.525 | 0.157 | 0.218 to 0.832 | **<0.001** |
| Religion: Muslim | -0.112 | 0.140 | -0.386 to 0.162 | 0.422 |  |  |  |  |
| Residence: Kenema | 3.435 | 0.199 | 3.045 to 3.826 | **<0.001** | 3.215 | 0.212 | 2.800 to 3.630 | **<0.001** |
| Time since HIV diagnosis | -0.083 | 0.017 | -0.117 to -0.049 | **<0.001** | -0.008 | 0.020 | -0.048 to 0.032 | 0.706 |
| Family/friend with HIV: yes | -1.328 | 0.172 | -1.664 to -0.991 | **<0.001** | -0.499 | 0.182 | -0.855 to -0.143 | **0.006** |
| Disclosed HIV status: yes | -1.172 | 0.149 | -1.463 to -0.881 | **<0.001** | -0.710 | 0.162 | -1.027 to -0.393 | **<0.001** |
| Enacted stigma: yes | 0.198 | 0.338 | -0.012 to 0.408 | 0.065 | 0.804 | 0.116 | 0.576 to 1.032 | **<0.001** |

### Associations between perceived and enacted HIV stigma domains

Given that enacted stigma was a predictor of perceived HIV stigma, we further explored the correlation between indicators of enacted sigma and the domains of perceived HIV stigma (Table 5). There was a weak positive correlation between perceived HIV stigma and enacted stigma (r=0.082, p = 0.041). Family isolation positively correlated with overall perceived stigma (r = 0.181, p < 0.001) and three of the four domains of perceived stigma. Across domains, family isolation moderately correlated with personalized stigma (r = 0.223, p < 0.001).

**Table 5.**
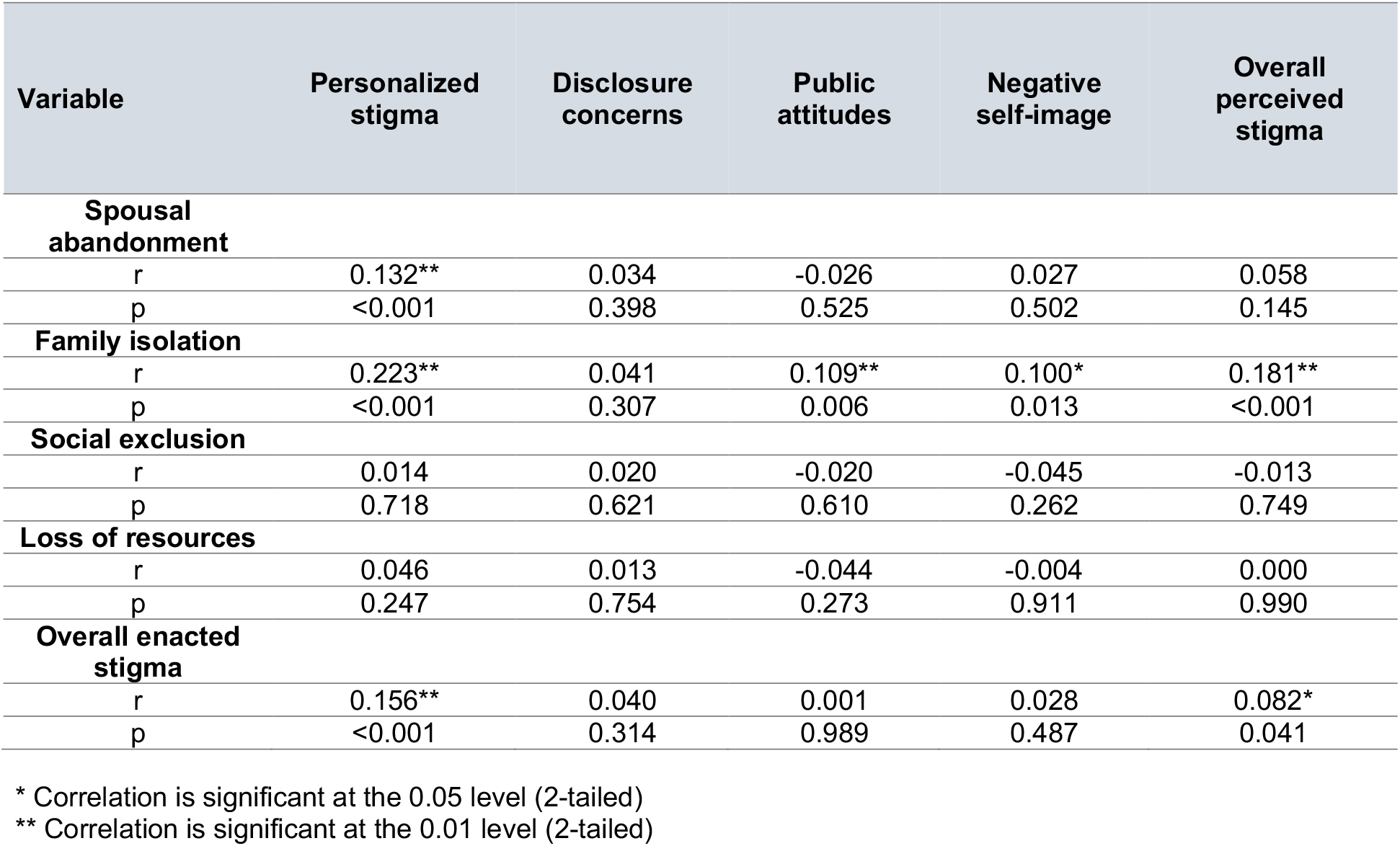
The Pearson correlation of the subscales of perceived and enacted HIV stigma

| Variable | Personalized stigma | Disclosure concerns | Public attitudes | Negative self-image | Overall perceived stigma |
| --- | --- | --- | --- | --- | --- |
| **Spousal abandonment** |  |  |  |  |  |
| r | 0.132** | 0.034 | -0.026 | 0.027 | 0.058 |
| p | <0.001 | 0.398 | 0.525 | 0.502 | 0.145 |
| **Family isolation** |  |  |  |  |  |
| r | 0.223** | 0.041 | 0.109** | 0.100* | 0.181** |
| p | <0.001 | 0.307 | 0.006 | 0.013 | <0.001 |
| **Social exclusion** |  |  |  |  |  |
| r | 0.014 | 0.020 | -0.020 | -0.045 | -0.013 |
| p | 0.718 | 0.621 | 0.610 | 0.262 | 0.749 |
| **Loss of resources** |  |  |  |  |  |
| r | 0.046 | 0.013 | -0.044 | -0.004 | 0.000 |
| p | 0.247 | 0.754 | 0.273 | 0.911 | 0.990 |
| **Overall enacted stigma** |  |  |  |  |  |
| r | 0.156** | 0.040 | 0.001 | 0.028 | 0.082* |
| p | <0.001 | 0.314 | 0.989 | 0.487 | 0.041 |

\* Correlation is significant at the 0.05 level (2-tailed)
\*\* Correlation is significant at the 0.01 level (2-tailed)

## DISCUSSION

This study aimed to adapt and validate a concise HIV stigma scale and assess perceived and enacted HIV stigma among PWH in Eastern and Southern Sierra Leone. We developed a 6-item HIV stigma scale with acceptable internal consistency. Exploratory factor analysis generated a four-factor scale capturing the dimensions of stigma in the following order of decreasing psychometric reliability: concern with public attitudes, personalized stigma, negative self-image, and disclosure concerns. The shortened scale offers a practical and efficient means of assessing stigma, which can be valuable in clinical and research settings. Furthermore, translating the scale into Krio, the national language of Sierra Leone, helped ensure accessibility for PWH with limited English proficiency.

The findings of our study shed light on the levels of perceived and enacted HIV stigma among PWH in Eastern and Southern Sierra Leone. The prevalence of perceived HIV stigma was high, with over two-thirds (68.6%) of participants reporting experiencing stigma. This aligns with previous studies that have demonstrated the pervasive nature of HIV stigma and its detrimental effects on the well-being and health of PWH [7-17]. On the other hand, the prevalence of enacted stigma was relatively low (6.7%) in this setting, which contrasts with findings from previous studies in sub-Saharan Africa, which found high levels of enacted stigma towards PWH [28-30]. The lower prevalence of enacted stigma in Sierra Leone could be attributed to the efforts made in recent years to reduce discrimination against PWH in the country, including awareness campaigns and community education programs [31]. However, as shown by our findings, even low levels of enacted stigma can have significant negative impacts on PWH, such as abandonment by spouse/partner and family isolation, emphasizing the critical importance of continued efforts to combat stigma and discrimination directed towards PWH in all its forms.

Furthermore, our study identified several factors associated with perceived HIV stigma. Higher levels of perceived HIV stigma were significantly associated with employment status, residing in Kenema (Eastern Province), and experiencing enacted stigma. Conversely, protective factors against perceived HIV stigma included a longer duration since HIV diagnosis, having a family member or friend with HIV, and disclosing HIV status. These findings are in agreement with previous studies which have highlighted the important role of psychosocial support and status disclosure in reducing HIV stigma [32, 33]. Similarly, the association between employment status and higher levels of perceived stigma has been documented. Additionally, the finding that residence in a specific region (Kenema) was associated with higher levels of perceived stigma is consistent with studies that observed regional variations in stigma attitudes and awareness [7].

The correlation analysis between the perceived and enacted stigma subscales further supports the intersectional nature of HIV stigma in Sierra Leone and other settings. Our finding of a weak positive correlation between total perceived HIV stigma and enacted stigma is consistent with studies that have highlighted the influence of enacted stigma on individuals’ perception of stigma [9]. Personalized stigma and family isolation showed significant positive correlations with all domains of perceived stigma, indicating that individuals who experienced spousal abandonment and family isolation were more likely to perceive higher levels of stigma across multiple dimensions. However, social exclusion and loss of resources did not demonstrate significant associations with perceived stigma. In terms of enacted stigma, only personalized stigma showed a significant positive correlation. These findings suggest that personal experiences of stigma, such as spousal abandonment and family isolation, have a stronger impact on perceived HIV stigma compared to social exclusion and loss of resources. Understanding these domains can inform interventions and support strategies to address specific areas of stigma and their implications for the well-being of PWH.

While our study provides valuable insights into perceived and enacted HIV stigma in Sierra Leone, several limitations should be considered. Firstly, the study was conducted in two specific geographic regions of Sierra Leone, which may limit the generalizability of the findings to other settings within Sierra Leone or other African countries. Additionally, the study relied on self-reported measures, which are subject to social desirability bias and may not capture the full extent of stigma experienced by individuals. Furthermore, the cross-sectional design of the study does not allow for causal inferences. It is also important to acknowledge the potential for recall bias, as participants may have had difficulty accurately recalling past experiences of stigma. Lastly, the study did not explore other potential factors that could influence perceived and enacted HIV stigma, such as socio-economic status, cultural beliefs, and access to healthcare services. Despite these limitations, our study contributes to the understanding of HIV stigma in Sierra Leone and provides a foundation for future research and interventions in this context.

## CONCLUSION

In summary, the adaptation and validation of the 6-item HIV Stigma Scale in the local context provides a concise and reliable tool for measuring perceived HIV stigma among PWH in Sierra Leone. The findings from our study have important implications for the design of HIV prevention, care, and psychosocial support programs in Sierra Leone. The high prevalence of perceived stigma underscores the need for targeted interventions that address societal attitudes, reduce discrimination, and promote inclusivity. Community-based education programs that challenge HIV-related stereotypes and promote accurate knowledge can help dispel misconceptions and reduce stigma. Additionally, efforts to enhance psychosocial support networks and encourage HIV status disclosure can help create a supportive environment for PWH and contribute to reducing stigma.

## Data Availability

The data presented in this study are available from the corresponding author upon reasonable request.

## AUTHOR CONTRIBUTIONS

GAY, LSB and RAS conceptualized and designed the study. EJS, RAK, PBJ, SAY, PEC, LMB, SPM and MG contributed to study concept and design. EJS, RAS, LSB collected the data. GAY conducted the statistical analysis. PBJ, PO, and SL contributed important intellectual content. All authors contributed to the interpretation of the data. GAY wrote the initial manuscript draft. All authors critically revised and approved of the final version. GAY is acting as the guarantor of this manuscript.

## ACKLOWLEDGEMENTS

We wish to acknowledge the staff at Bo and Kenema Government Hospitals and people living with HIV without whose help this study would not have been successful.

## ETHICAL APPROVAL

Ethical approval was obtained from the Njala University Institutional Review Board (approval date 11 June 2022).

