## Supplemental Material 1 for "Perceived and Enacted HIV-Related Stigma in Eastern and Southern Sierra Leone: A Psychometric Study of an HIV Stigma Scale"

**Form No……………**

***INSTRUCTIONS*:** This survey is anonymous; your participation is strictly voluntary.

Your identity or health information will not be revealed.

By answering these questions, your consent to participate in this study is implied

Please write or circle the answers you believe to be accurate. There are NO right or wrong answers

**Age** *(years) .……………….*

**Gender**	☐Male	☐Female

**Relationship status**	☐Single ☐Married ☐Widowed ☐Divorced

**Highest level of education** ☐None ☐Primary ☐Secondary ☐College/tertiary

**Employment** ………………………………………………………………………

**Monthly income/earning (Leones)** ……………………………………………

**Religion** ☐Christian ☐Muslim ☐Other……………………………………...

**HIV status** ☐Positive ☐Negative

**How long have you had the diagnosis (years**)?...............................................

**Have you ever disclosed your status to someone?**

Yes	No

**Are you taking medications or treatment for your infection?**

Yes	No

**Have you felt abandoned by spouse or partner?**

Yes	No

**Have you ever felt isolated by family members?**

Yes	No

**Have you ever felt excluded from social activities?**

Yes	No

**Do you think your illness has affected your career progression?**

Yes	No

*PLEASE ANSWER ALL 12 QUESTIONS*

**PERSONALISED**

1. Some people avoid touching me once they know I have HIV

| Strongly disagree  1 | Disagree  2 | Agree  3 | Strongly agree  4 |
| --- | --- | --- | --- |

2. People I care about stopped talking to me after learning I have HIV

| Strongly disagree  1 | Disagree  2 | Agree  3 | Strongly agree  4 |
| --- | --- | --- | --- |

3. I have lost friends by telling them I have HIV

| Strongly disagree  1 | Disagree  2 | Agree  3 | Strongly agree  4 |
| --- | --- | --- | --- |

**DISCLOSURE CONCERNS**

4. Telling someone I have HIV is risky

| Strongly disagree  1 | Disagree  2 | Agree  3 | Strongly agree  4 |
| --- | --- | --- | --- |

5. I work hard to keep my HIV a secret

| Strongly disagree  1 | Disagree  2 | Agree  3 | Strongly agree  4 |
| --- | --- | --- | --- |

6. I am very careful who I tell that I have HIV

| Strongly disagree  1 | Disagree  2 | Agree  3 | Strongly agree  4 |
| --- | --- | --- | --- |

**ENACTED STIGMA/PUBLIC ATTITUDES**

7. People with HIV are treated like outcasts or rejects

| Strongly disagree  1 | Disagree  2 | Agree  3 | Strongly agree  4 |
| --- | --- | --- | --- |

8. Most people believe a person who has HIV is dirty

| Strongly disagree  1 | Disagree  2 | Agree  3 | Strongly agree  4 |
| --- | --- | --- | --- |

9. Most people are uncomfortable around someone with HIV

| Strongly disagree  1 | Disagree  2 | Agree  3 | Strongly agree  4 |
| --- | --- | --- | --- |

**NEGATIVE SELF IMAGE**

10. I feel guilty because I have HIV

| Strongly disagree  1 | Disagree  2 | Agree  3 | Strongly agree  4 |
| --- | --- | --- | --- |

11. People’s attitudes about HIV make me feel worse about myself

| Strongly disagree  1 | Disagree  2 | Agree  3 | Strongly agree  4 |
| --- | --- | --- | --- |

12. I feel I’m not as good a person because I have HIV

| Strongly disagree  1 | Disagree  2 | Agree  3 | Strongly agree  4 |
| --- | --- | --- | --- |
